# Modelling and Preliminary Clinical Validation of Home-based Menstrual Neuromodulation Therapy

**DOI:** 10.1101/2024.02.02.24302224

**Authors:** Emilė Radytė, Laura Stankevičiūtė, Ervinas Bernatavičius, Alexander Cook, Yvinna Tamiris Rodrigues, Tatiana Camila Lima de Alves Silva, Maria Thereza Albuquerque Barbosa Cabral Micussi, Rodrigo Pegado

## Abstract

Primary dysmenorrhea (PD), characterised by chronic pelvic pain during menstruation, significantly impairs the quality of life for many women. This paper presents a modelling and clinical validation study of the novel Nettle™ device for at-home transcranial direct current stimulation (tDCS) for alleviating PD symptoms. Specifically, we aimed to investigate the electric field patterns induced by Nettle™ and its immediate efficacy in reducing menstrual pain and improving functionality. Finite element method (FEM) simulations, using realistic head models, assessed the electric field distribution, targeting key brain regions involved in pain processing. A single-centre triple-blinded, sham-controlled study involving 34 women was conducted to compare the effects of active and sham tDCS. Results demonstrated a clinically meaningful decline in menstrual pain symptoms in the active group, with medium effect sizes for both pain reduction (Cohen’s d=0.53) and functionality (Cohen’s d=0.47), based on Nettle™’s protocol focused on the electric field within the medial prefrontal cortex. Limitations include the use of generalised brain models and small sample size, highlighting the need for further research with comprehensive modelling and larger clinical trials to validate and understand the effects of Nettle™ as a menstrual neuromodulation therapy.

**Clinical Relevance:** This study underscores Nettle™’s potential as a non-invasive, cost-efficient intervention for PD, with implications for broader applications in women’s health.

## I. Introduction

Dysmenorrhea, defined as chronic, recurring pelvic pain during menstruation, manifests predominantly as primary dysmenorrhea (PD) or less commonly as secondary dysmenorrhea. PD, a prevalent gynaecological disorder absent of other pathology, induces pain preceding and during menses. It is often accompanied by psychological symptoms—such as anxiety and depression—along with physiological effects like nausea, migraines, and diarrhoea. The collection of these symptoms is referred to as premenstrual syndrome (PMS). An estimated 17-90% of women of reproductive age experience menstrual symptoms, which remain an inadequately addressed gynaecologic issue [1]. These symptoms can significantly impact daily functioning and quality of life, particularly at the onset of the menstrual period, affecting educational and occupational activities.

PD is linked to alterations in brain metabolism and functional connectivity in regions implicated in pain modulation, including the primary motor cortex (M1), medial prefrontal cortex (mPFC), posterior cingulate cortex, and the insula, which exhibit abnormal functional patterns [2]. These areas contribute to the emotional and affective regulation of pain, cognitive control, and pain generalisation. Women with PD demonstrate heightened theta wave (4-7 Hz) activity in these pain-associated cerebral regions [3]. Similarly, women with severe premenstrual symptoms demonstrate alpha (8-12 Hz) wave asymmetry in the prefrontal regions of the brain [4]. The interrelation of pain with depressive and anxious states may be crucial for elucidating the role of theta and alpha activity in the sensory-emotional pain processing specific to PD. Crucially, these neurocognitive alterations are not confined to menstrual episodes, suggesting PD should be understood as a chronic pain condition [5]. This understanding of the enduring neurocognitive and behavioural changes in women with PD paves the way for novel, non-pharmacological treatment approaches that are cost-effective and have minimal side effects.

Non-invasive brain stimulation (NIBS) has been used in several clinical conditions aiming to improve motor, cognitive, affective-behavioural and physical function. The clinical application of NIBS has been gaining prominence in new consensuses and guidelines in physical and cognitive rehabilitation aimed to improve or restore function in chronic diseases. Transcranial direct current stimulation (tDCS) is particularly noted for its scalability and involves administering a constant low-intensity microcurrent of 1 to 4 mA to the scalp. This results in modulated cortical excitability, contingent on specific parameters such as duration, intensity, and electrode placement, which contribute to its therapeutic effects.

Several investigations have been published, ranging from computational simulations to animal experiments and human clinical trials, supporting the use of tDCS as an effective, non-invasive, and cost-efficient intervention with minimal side effects for disorders such as major depressive disorder, fibromyalgia, and stroke rehabilitation, among others [6]. In particular, studies examining tDCS in women with PD have reported beneficial outcomes for pain management [7], functionality and anxiety [8]. Importantly, the efficacy of tDCS is significantly influenced by the configuration of electrode placement. Anodic stimulation at the primary motor cortex (M1), using a C3/Fp2 montage as per the 10/20 EEG system, enhances pain modulation, while similar stimulation at the dorsolateral prefrontal cortex (DLPFC), using an F3/Fp2 montage, has been associated with reduced anxiety.

tDCS is typically prescribed by physicians or physiotherapists, varying with national regulations, and administered under professional supervision in clinical settings. The push for remote rehabilitation, including tDCS, represents a significant shift in patient care, a trend accelerated by the COVID-19 pandemic’s social distancing measures. While in-person treatment at research facilities or clinics is effective, it presents logistical challenges, requiring patients to commit to numerous sessions. This commitment often incurs considerable costs for transportation, time, and meals, and can lead to absence from work or school, limiting overall treatment accessibility.

A remote tDCS model that is safe, user-friendly, and associated with low adverse effects could enhance adherence and provide an alternative therapy for women suffering from primary dysmenorrhea (PD) and PMS [9]. Existing remote tDCS protocols have targeted chronic pain and various musculoskeletal and behavioural disorders but have not specifically addressed women’s health concerns, including PD.

Nettle™ by Samphire Neuroscience, a novel home-use tDCS device, has been developed to meet this need. It operates based on validated stimulation parameters and electrode configurations to mitigate PD symptoms. Designed to resemble a women’s hair accessory for discretion, Nettle™ is a wireless, Bluetooth-enabled wearable device managed through the Samphire App. The app guides users through session setup, checks for contraindications, customises session parameters, and allows for session control, including starting, pausing, and stopping, as well as troubleshooting support and collection of post-session feedback.

This is a modelling and clinical validation study with healthy volunteers aiming to evaluate the potential of the Nettle™ home-based tDCS system with a focus on women’s health. The specific aims of this study were to (1) understand the electric field patterns and strengths associated with the use of Nettle™, and (2) assess the immediate efficacy of Nettle™ in alleviating pain and mood symptoms associated with menstruation.

## II. METHODS

### A. Electric Field Modelling & Simulations

Head models for tDCS simulations were created using the finite element method (FEM) with a realistic head model mesh derived from an example subject’s structural magnetic resonance scans in SimNIBS [10]. Default tissue conductivities were used (bone (0.010 S/m), scalp (0.465 S/m), grey matter (0.275 S/m), white matter (0.126 S/m), cerebrospinal fluid (1.654 S/m)). Simulation results were visualised with Gmsh and MATLAB, and all calculations were conducted in MATLAB. Active electrode montages were determined by simulating electrodes on a 10-20 EEG cap that represents the potential positions of electrodes on the scalp and compiles a matrix of the electric fields generated by every electrode while keeping the return electrode constant, then optimised to reach a particular field strength at the target. DLPFC MNI coordinates were (−49.33; 52.25; 73.03), and M1 coordinates were (−64.32, -15.38, 82.66) all in millimetres.

For tDCS modelling, rectangular 5.0 cm x 2.0 cm sponge electrodes were used with a 2.5 mm thickness. The specific positions in the 10-20 electrode system for each of the montages are shown in Figure 1.

**Fig. 1.**
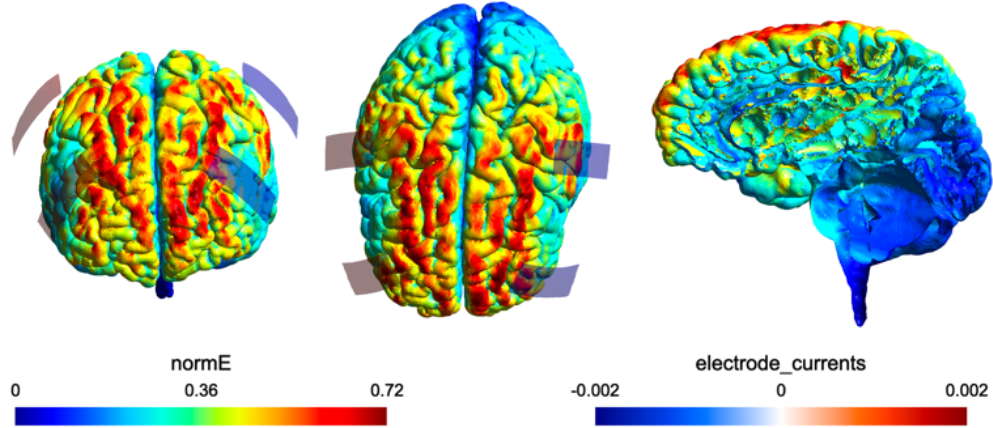
Anterior, dorsal and sagittal views of electric field modelling results for the Nettle™ device. Electric field modelling results suggest that under Nettle™’s stimulation protocol, the majority of the electric field strength (measured in V/m units, max at 0.72 V/m) is concentrated in the dorsolateral and medial prefrontal cortices, as well as the motor cortex. It also shows that there is negligible direct penetration of the electrical stimulation into subcortical areas of the brain. Electrode currents are measured in A.

### B. Preliminary Clinical Validation Trial Design

The clinical validation study was a single-centre triple-blinded study conducted at the Federal University of Rio Grande do Norte (UFRN) in Natal, Brazil. The study was previously approved by the Research Ethics Committee of UFRN under number 5.508.364. Inclusion criteria admitted participants, who: (1) were 18-45 years old, (2) had a regular menstrual cycle, (3) were not lactating, (4) had no history of brain surgery, tumours, or intracranial metal implantation, and (5) had no history of chronic genitourinary infections, alcohol, or drug abuse. Exclusion criteria were: (1) patients presenting with a history of dizziness or epileptic disease, (2) pregnancy, and (3) metal implants in the head. A total of 34 women participated in the study. Participants were enrolled by the blinded investigators and randomly allocated (1:1) to receive active tDCS (n = 18) or sham tDCS (n = 16). Stratified randomisation was done using the order of entry into the study to assign each participant to either the active or sham group. An external blinded research assistant generated the allocation sequence. Participants and evaluators were blinded to group allocation throughout the trial. The study was run for a single intervention menstrual cycle in order to establish the clinical trend effects of the intervention.

### C. Menstrual Neuromodulation Therapy Intervention

The study consisted of a single-day assessment and intervention with 4 stages: (1) initial meeting with the participants to introduce the project, complete informed consent procedures and baseline evaluation of menstrual symptoms, (2) 5-day use of Nettle™ at-home by the participant in the 5 days of their late luteal phase, (3) intervention evaluation on the first day of their menstrual cycle following the use of Nettle™, (4) evaluation of menstrual symptoms one month following the use of Nettle™.

The Samphire App was configured in the participants’ native (Brazilian Portuguese) language to ensure accessibility, with an option to change it to English.

Nettle™ was set to deliver direct current to the scalp at the M1 area of the brain (M1, anodal EEG 10-20 C3/C4). The device has four sponge electrodes (10.25 cm^2^) that must be impregnated by the user with a saline (0.9% NaCl) solution before placing the device on the head. Each participant self-administered five 20-minute sessions, one daily, in their late luteal phase.

In the active tDCS group, the current was ramped up from 0 to 2.0 mA over 30 seconds, held constant for 19 minutes, and ramped down over 30 seconds, for a total session time of 20 minutes. In the sham tDCS group, to ensure blinding, the current was ramped up to 2 mA over 30 seconds, then ramped down to 0.01 mA, considered negligible, and held at the negligible output for 19.5 minutes, for a total session duration of 20 minutes. This method produces the same mimic sensations such as itching, and tingling observed during active tDCS, enabling blinding to be achieved [9], as shown in a previous usability study. Both active and sham tDCS groups used identical devices, with the session type being determined by a previously randomised code input into the Samphire App. During tDCS use, participants were instructed to feel free and continue with their normal routine.

### D. Outcome measures

Primary outcomes were changes in pain scores on a visual analog scale and the 6-minute walking test assessing participant functionality. Secondary measures included a range of tests to assess mental health impacts, such as low mood, anxiety, and negative and positive emotionality, which are beyond the scope of this paper.

### E. Statistical analysis

SPSS software version 19.0 (IBM Corp., Armonk, NY, USA) was used for statistical analyses. Clinical and sociodemographic characteristics were described by means and standard deviations or by frequency tables for qualitative parameters. A chi-squared test was used to compare the distributions of qualitative variables. The Shapiro-Wilk test was used to assess the normality of the distribution. To compare data between groups, an unpaired t-test or Mann-Whitney were used. Statistical significance was set at p < 0.05.

## III. RESULTS & DISCUSSION

### A. Electrical Field Modelling

Electric field modelling results suggested that under Nettle™’s stimulation protocol, the majority of the electric field strength is concentrated in the mPFC, with secondary locations in the DLPFC and motor cortices) (Figure 1). Recent evidence suggested strong connections between the mPFC and the posterior cingulate and insula [11], which may explain why even after a single cycle use, users could expect to experience clinically significant results.

### B. Effect on pain perception within a single cycle

In line with observations from the NIBS literature [12], there was a large initial sham and active effect from baseline to the intervention menstrual cycle, associated with the novelty of treatment. However, according to the predictions around the longer-term neuroplasticity effects of NIBS on pain perception [13], symptoms rebounded in the sham group in the follow-up menstrual cycle, while continuing to be suppressed in the active group (average pain severity decrease of -52.8% in the active group vs -24.43% decrease in the sham group). There was a medium effect size (*Cohen’s d=0*.*53*) (Figure 2). This change suggests a clinically meaningful decline in menstrual pain symptoms in the active group after a single cycle of use.

**Fig. 2.**
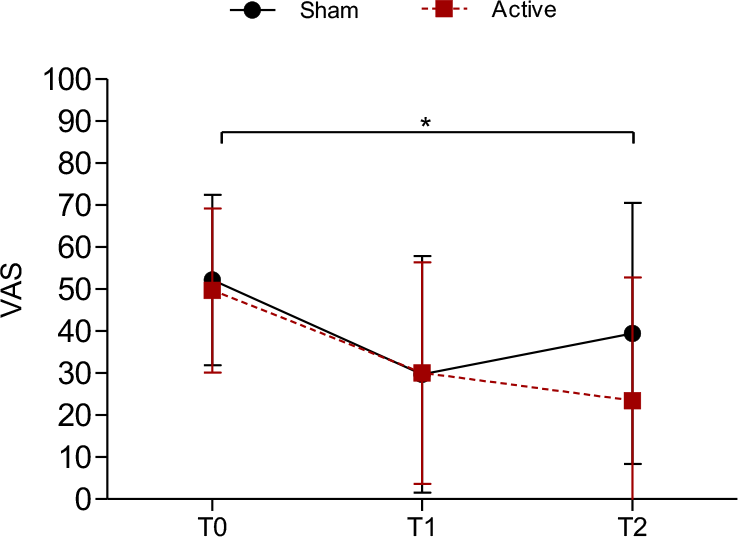
Pain Visual Analog Scale measure for active and sham groups. A significant difference was found between T0 and T2 (p = 0.002, *d =* 0.53 (CI - 1.215; 0.155) for the active, but not the sham group. *Denotes significant intragroup difference.

### C. Effect on functionality within a single cycle

There were temporary increases in functionality, or physical performance, in the active but not the sham group after a single cycle of use, with a medium effect size (*Cohen’s d=0*.*47*) (Figure 3).

**Fig. 3.**
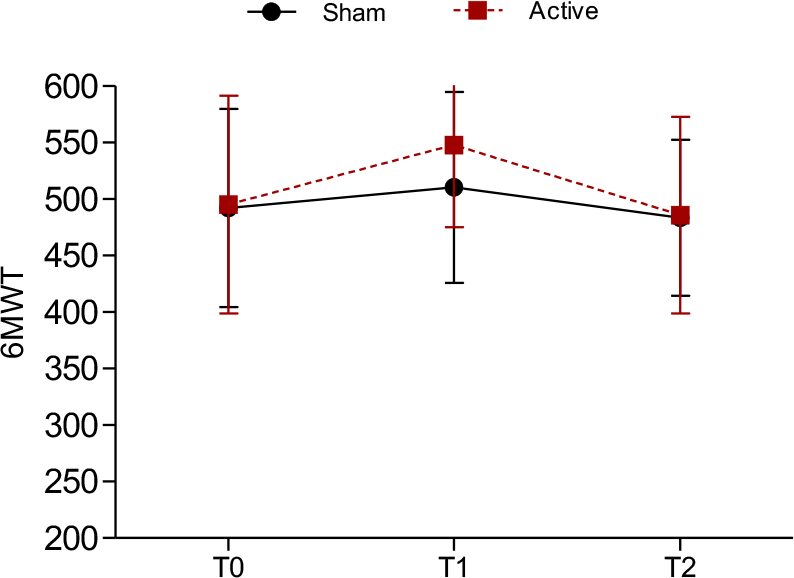
Six-minute walking test (6MWT) functionality measure for active and sham groups. A significant difference was found between T0 and T1 (p = 0.04, *d =* 0.47 (CI -0.20; 1.15)) for the active, but not the sham group.

This is expected, as physical effects are usually considered to be a short-rather than longer-term effect of NIBS underlying this menstrual neuromodulation therapy.

## IV. LIMITATIONS & CONCLUSION

This paper presents a brief overview of the reasoning, modelling, and preliminary clinical and public health implications of a new device, Nettle™, able to deliver non-invasive menstrual neuromodulation therapy, with two core limitations.

Firstly, the brain models used in this study are not derived from a specific participant in the study but are rather a simplified representation of the brain morphology and tissue behaviour based on population data. This type of electric field modelling does not account for the temporal dimension of the electric field magnitude or the spatial dimension and morphology of individual neurons. A multi-compartmental neuronal model, able to account for the orientation of electric field vectors, the activating function and network dynamics would all result in a more accurate model of the amplitudes and dispersion of electrical fields in a specific user.

Secondly, this was a preliminary clinical study, the purpose of which was to validate the clinical premise of using NIBS in the management of long-term, chronic conditions disproportionately impacting the quality of life of women. Thereby, it is likely to have been underpowered from a sample size and a duration perspective to be able to draw more conclusive results on the effect, variability and effect size of the clinical efficacy of the Nettle™ device.

Further studies are set to explore more complex modelling and simulations of the underlying biophysical effects of Nettle™ on individual neuronal and network connectivity, as well as larger clinical samples for more conclusive results.

Overall, this paper demonstrates that NIBS could be an effective modality for the management of long-term conditions in the area of women’s health, especially for menstrual symptoms, such as premenstrual syndrome and menstrual pain.

## Data Availability

All data produced in the present study are available upon reasonable request to the authors.

